# Bidimensional Perfectionism and Psychological Distress: The Roles of Self-Esteem and Self-Compassion

**DOI:** 10.1101/2024.07.24.24310699

**Authors:** Cheuk Hei Peony Chung, Antoinette Marie Lee

**Author notes:** **CRediT Author statement:** **Cheuk Hei Peony Chung:** Conceptualization, Methodology, Formal analyses, Investigation, Data Curation, Writing-Original Draft, Visualization **Antoinette Marie Lee:** Conceptualization, Methodology, Writing-Review & Editing, Supervision.

## Abstract

Perfectionism as a personality trait can be seen as having both adaptive and maladaptive dimensions. Nevertheless, their relationships with psychological distress remain mixed in the current literature. Previous studies were also limited by the use of impure measurements and the failure to statistically control for the effects of the other dimension. By addressing these major limitations and exploring the mediating and moderating roles of self-esteem and self-compassion, the current study provides an in-depth examination of the relationships between bidimensional perfectionism and psychological distress. In a community sample of 503 adults, results supported a bidimensional view of perfectionism, with maladaptive perfectionism positively predicting psychological distress and adaptive perfectionism being unrelated to psychological distress. Self-esteem was found to mediate the relationships between both dimensions of perfectionism and psychological distress. Self-compassion was only found to moderate the relationship between maladaptive perfectionism and self-esteem. Whilst the maladaptive nature of maladaptive perfectionism was supported in this study, findings suggested that adaptive perfectionism remains a more complicated construct. Future studies should aim at clarifying the nature and psychological outcomes of adaptive perfectionism.

## Literature Review

### Perfectionism as a Bidimensional Construct

Perfectionism—the striving for flawlessness, can be seen as having both adaptive and maladaptive dimensions. The current literature has been dominated by research on maladaptive perfectionism, a trait that can be defined as having high personal standards and tendencies to be self-critical in self-evaluations (Rice & Stuart, 2010). It is not until the recent two decades that the adaptive dimension is also receiving more attention. Whilst also showing the striving for high standards, adaptive perfectionism is a personality trait that enables one to derive satisfaction from achievements, whilst remaining tolerant without resorting to harsh self-criticism when imperfections occur (Stoltz & Ashby, 2007).

The dual process model (Slade & Owens, 1988) could be used to understand the two dimensions of perfectionism, based on their underlying functional differences. On the basis of Skinner’s (1938) behavioural theory, ‘negative’ (maladaptive) perfectionism is driven by negative reinforcement—maladaptive perfectionism is developed and maintained through the avoidance of negative consequences, including the fear of failure and disapproval from others. Contrarily, ‘positive’ (adaptive) perfectionism is believed to be driven by positive reinforcement, where individuals are fueled by the pursuit of success, excellence, approval, and a sense of accomplishment, etc.

Empirical evidence supports a bidimensional view of perfectionism. Confirmatory factor analysis of the major multidimensional perfectionism scales has revealed two dimensions of perfectionism: ‘perfectionistic concerns’ and ‘perfectionistic strivings’ (Frost et al., 1993; Slade and Owens, 1998). Whilst ‘perfectionistic concerns’ were consistently found to be linked with negative psychological outcomes, ‘perfectionistic strivings’ was found to be linked with positive psychological outcomes (Stoeber, 2017).

### Maladaptive Perfectionism

Traditionally, perfectionism was studied through a psychopathological lens (Stoeber & Otto, 2006). Psychodynamic theorists stress that perfectionism is a sign of a neurotic and disordered personality (Burns, 1980; Hollender, 1965; Horney, 1951; Missildine, 1963), giving rise to psychological distress and psychopathologies, with the most common being depression and anxiety.

#### Perceived Discrepancy Between Standards and Performance

Perceived discrepancy, defined as ‘the perception that one consistently fails to meet the high standards one has set for oneself’ (Slaney et al., 2002, p. 69), is one of the central and defining features of maladaptive perfectionism. Such a feature is considered maladaptive since worrying about achieving high standards often leads to anxiety, and ‘failures’ to achieve one’s unrealistically high standards often results in intense self-criticism and feelings of worthlessness (Slaney et al., 2001). Moreover, maladaptive perfectionism is linked with the tendency to reappraise standards to higher levels upon reaching them, with the belief that they were insufficiently demanding (Shafran et al., 2002). This increases the difficulty in succeeding and the likelihood that one will experience perceived failure, forming a negative cycle of self-criticism and dissatisfaction.

#### Cognitive Distortions

Maladaptive perfectionism is also related to patterns of ‘cognitive distortions’— inaccurate and inflexible interpretations of information that cause and maintain negative bias and beliefs about oneself and the world (Beck, 1976). One common distortion is ‘all-or-nothing’ thinking, where individuals evaluate their experience in a dichotomous and inflexible manner, perceiving outcomes as either ‘all right’ or ‘all wrong’. Individuals high in maladaptive perfectionism often believe they are failures if they cannot reach ‘perfection’, failing to see the continuum between the two extremes. Since their standards are often too high to achieve, they are always left thinking that they are ‘second-rate losers’ (Burns, 1980).

These individuals are also often guided by ‘should’ statements, so much so that psychoanalytic theorist Karen Horney (1951) describes ‘neurotic perfectionism’ as the ‘tyranny of the shoulds’. ‘Should’ statements reflect rigid and harsh self-talk, e.g., telling oneself *‘I should always work hard’* or *‘I shouldn’t have made any mistakes’*. Irrational ‘should’ statements imply one has to be perfect, all-knowing, or all-powerful, which creates an inappropriate sense of guilt as they do not represent sensible standards (Burns, 1980, p.202). Instead of treating oneself kindly with self-acceptance, ‘should’ statements cause one to become ‘trapped by nonproductive self-critical ruminations’ when facing mistakes, leading to unrealistic negative self-image and depression (Burns, 1980, p. 38).

The cognitive distortion of ‘selective attention’ also serves to maintain individuals’ maladaptive perfectionism (Hollender, 1965). Like an ‘inspector at the end of a production line’, individuals with maladaptive perfectionism are ‘constantly on the alert for what is wrong’ but ‘seldom focuses on what is right’ (Hollender, 1965, p.95). They selectively attend to experiences of failures, even minor mistakes, while ignoring and discrediting instances of success. They also tend to attribute their success to external sources, contributing to their lowered perceived self-efficacy (Lo & Abbott, 2013).

### Adaptive Perfectionism

Hamachek (1978) was amongst of the first to propose the concept of ‘normal perfectionism’, as opposed to ‘neurotic perfectionism’. Through clinical observations, Hamachek (1978) described individuals with normal perfectionism as ‘skilled artists’ who derive a ‘real sense of pleasure from the labour of a painstaking effort’, those ‘who feel free to be less precise as the situation permits’ (p.26), and is motivated by a ‘desire for improvement’ (p.28).

Rather than being driven by the fear of failure, adaptive perfectionism is fueled by the motivation to succeed and improve (Hamachek 1978; Stoeber & Otto, 2006). With such a trait, individuals strive for high-level goals due to their motivation to pursue success, excellence, approval, and a sense of accomplishment (Slade & Owens, 1988). Instead of worrying about one’s deficiencies and what could go wrong, adaptive perfectionism enables individuals to ‘focus on their strengths and concentrate on how to do things right’ (Hamachek, 1978, p.28).

Adaptive perfectionism also entails the ability to feel satisfied and accomplished upon reaching one’s high standards (Missildine, 1963). It allows individuals to reap the fruits of their strivings, leading to good feelings about themselves and their work. This is contrasted with maladaptive perfectionism, which often brings about dissatisfaction even when goals are achieved, as manifested as constant standards resetting.

Moreover, with regards to experiencing failures and mistakes, adaptive perfectionism is not related to the harsh self-criticism, self-defeating and ruminative behaviours which characterise maladaptive perfectionism (Bieling et al., 2003; Enns et al., 2001; Stoltz & Ashby, 2007). Adaptive perfectionism enables one to be accepting of occasional mistakes by being more realistic and flexible, as well as to adopt practical coping strategies (Li et al., 2015). Hence, individuals high in this trait are less likely to overinterpret their occasional mistakes as reflecting their unworthiness, but instead allow themselves to be ‘less precise’ sometimes, while maintaining their motivation to strive for high standards.

#### Perfectionism and Psychological Distress: Empirical Evidence

Echoing theories, maladaptive perfectionism has been consistently found to be related to psychological distress. Cross-sectional studies showed that it is related to stress (Einstein et al., 2001), depression (Dunkley et al., 2000; Minarik & Ahrens, 1996; Sherry et al., 2014; Wang et al., 2009, Xie et al., 2019), anxiety (Anthony et al., 1998; Coles et al., 2003; Dunkley et al., 2000; Einstein et al., 2001), and general distress (Kong et al., 2021). Longitudinal studies also revealed that maladaptive perfectionism predicted increases in depressive symptoms in non-clinical samples (Smith et al., 2017), and the persistence of depressive symptoms in participants with depression (Enns & Cox, 2005; Hewitt et al., 1996). In one recent experiment, Hummel et al., (2023) experimentally induced maladaptive perfectionism and negative affect to test their possible bidirectional relationship. Results showed a unidirectional causal relationship from maladaptive perfectionism to negative affect, particularly in participants with high trait perfectionism.

Contrarily, although adaptive perfectionism has been consistently found to be linked to positive well-being (Stoeber & Otto, 2006), evidence regarding its relationship with psychological distress is less consistent. There is still debate on whether adaptive perfectionism per se is unproblematic, and whether it can protect one against psychopathology (Gäde et al., 2017; Stoeber & Otto, 2006). Some studies found that adaptive perfectionism is related to lower levels of psychological maladjustment, including depression (Abdollahi et al., 2020; Hill et al., 2004), burnout (Hill & Curran, 2016), and neuroticism (Costa & McCrae, 1990; Hill et al., 1997). Adaptive perfectionists as identified through cluster analyses were also found to show lower depression and anxiety symptoms than maladaptive perfectionists or non-perfectionists (Dixon et al., 2004; Mobley et al., 2005; Rice & Dellwo, 2002; Rice et al., 2003; Rice & Mirzadeh, 2000; Rice & Slaney, 2002).

Yet, other studies failed to find a relationship between adaptive perfectionism and psychological distress. It was found to be unrelated to depression (Bieling, 2004; Frost et al., 1993; Slaney et al., 2001; Rice et al., 1998; Wang et al., 2007; Wang & Zhang, 2017; Di Schiena et al., 2012), worry (Rice et al., 1998; Wang et al., 2007), and anxiety (Bieling 2004).

Furthermore, some studies have even found that adaptive perfectionism was linked with psychological maladjustment, including negative affect (Bieling et al., 2003; Dunkley et al., 2003), neuroticism (Cox et al., 2002; Enns et al., 2001), depression (Bieling et al., 2004; Cox et al., 2002; Lynd-Stevenson & Hearne, 1999), and anxiety (Bieling et al., 2004; Hill et al., 2004).

Such mixed results regarding the adaptive perfectionism and distress link could be due to a few reasons. Firstly, different measurements were used across studies. Most studies even used different scales to measure adaptive perfectionism in the same study. However, combining multiple scales that conceptualize adaptive perfectionism differently is believed to attenuate the connection between it and other variables (Ashby & Rice, 2002). Secondly, the two commonly used measurements of adaptive perfectionism: the ‘personal standards’ subscale from the Frost Multidimensional Perfectionism Scale (FMPS) (Frost et al., 1994) and the ‘self-oriented perfectionism’ subscale from the Multidimensional Perfectionism Scale (MPS) (Hewitt & Flett, 2004) were found to also measure dimensions of maladaptive perfectionism (DiBartolo et al., 2004; Owens & Slade, 2008), making them ‘impure’ measures (Ashby & Rice, 2002). Thirdly, all studies that found a lack of or negative relationship between adaptive perfectionism and psychological distress did not control for maladaptive perfectionism as a covariate. Since the two dimensions of perfectionism were consistently found to be correlated (Stoeber & Otto, 2006), i.e., individuals are likely to show both traits of adaptive and maladaptive perfectionism, the relationship between adaptive perfectionism and psychological distress is likely to be obscured by the effects of maladaptive perfectionism. Supporting this interpretation, studies that did not support the relationship between, or a showed positive relationship between adaptive perfectionism and negative mental health outcomes either became positive evidence, or null after a partial correlation was done (Stoeber & Otto, 2006). Lastly, there is possibly the existence of other variables (e.g., self-compassion) in understanding the adaptive perfectionism–distress link. However, studies that do this (e.g., moderation studies) are disproportionately focused on maladaptive perfectionism, limiting our understanding of adaptive perfectionism (Stoeber, 2017).

Hence, in exploring the relationship between adaptive perfectionism and psychological distress, the existing literature currently lacks studies that 1) adopt a consistent conceptualization of adaptive perfectionism, 2) measure such trait using a pure scale, 3) control the effects of maladaptive perfectionism, and 4) take into account the effects of other associated variables.

### Self-Esteem as a Mediator

Self-esteem is widely believed to play an integral role in the construct of perfectionism (Ashby & Rice, 2002). Preusser et al. (1994) even argued that depression is not, per se, the direct outcome of perfectionism, but mediated by self-esteem. Despite its integral role, literature on the mediating role of self-esteem remains mixed. Similarly, it is proposed here that this is due to the use of impure measurements and the failure to control for the effects of the other dimension. By studying self-esteem as a mediator whilst addressing these limitations, this study could further (1) fill the research gap on the mixed literature of self-esteem as a mediator, and (2) provide more insight into the relationship between bidimensional perfectionism and psychological distress.

Self-esteem is ‘the degree to which the qualities and characteristics contained in one’s self-concept are perceived to be positive’ (American Psychological Association, 2015). It involves a sense of self-worth, where healthy levels of self-esteem lead to one’s evaluation that one is worthy and likable (Rosenberg, 1965). It gives one ‘the feeling of being worthy, deserving, entitled to assert one’s needs and wants and to enjoy the fruits of one’s effort’ (Branden, 2021, p.8). Contrarily, low self-esteem is characterized by negative feelings towards the self and is linked with perceived low self-competencies. Individuals with low self-esteem typically exhibit less confidence, more negative self-talk, and self-doubt (Baumeister et al., 1989). Being a subjective measure, it does not necessarily reflect the actual competencies of an individual (Fearn et al., 2021). Whilst high self-esteem is linked with psychological well-being, was found to buffer negative psychological outcomes; low self-esteem was found to be linked with a plethora of psychological maladjustments, including depression and anxiety symptoms (Sowislo & Orth, 2013).

Individuals with high levels of maladaptive perfectionism tend to equate their self-worth with achievement and success (Burns, 1980; Shafran et al., 2002; Hewitt et al., 1996). Whilst an occasional achievement of ‘perfection’ leads to temporary positive feelings of themselves, they often struggle with feelings of failure and inadequacy (Burns, 1980). Since they do not allow slight leeway in making mistakes, coupled with their tendency to engage in dichotomous thinking, they are likely to perceive even the slightest mistakes as feedback that they have failed and are worthless as individuals (Hewitt et al., 1996). Furthermore, their tendency to selectively attend to failures also gives them a distorted and unrealistic view of their actual abilities, causing them to think lowly of themselves even if they are performing objectively well. Such patterns of constant self-dissatisfaction and criticism are believed to lead to a ‘precipitous loss in self-esteem that can trigger episodes of severe depression and anxiety’ (Burns, 1980, p.34).

Correlational studies support the relationship between maladaptive perfectionism and low self-esteem (Ashby & Rice, 2002; Rice et al., 1998; Slaney et al., 2001; Wang et al., 2007). However, mediation studies showed mixed evidence regarding the mediating role of self-esteem between maladaptive perfectionism and psychological distress. Some studies found the mediating role of self-esteem between maladaptive perfectionism and depression (Chai et al., 2020; Moroz & Dunkley, 2015; Preusser et al., 1994; Zhang & Cai, 2012), and anxiety amongst children (Chyug, 2012) and sports athletes (Mehrjoyan & Rahimi, 2021). Yet, such a mediating role was not found in relation to depression in Rice et al. (1998) and Park et al. (2020)’s studies, and was only found amongst male participants in relation to anxiety (Lasota & Kearney, 2017).

Contrarily, adaptive perfectionism is believed to lead to higher self-esteem (Hamachek, 1978; Missildine, 1963). Striving for perfection and excellence has always been considered healthy (Adler 1956; Maslow 1971, Murray 1938). When driven by the motivation to succeed and improve, it is thought to enhance one’s feelings of self-worth (Frost et al., 1993; Slade & Owens, 1988). Moreover, without engaging in cognitive distortions (e.g., dichotomous thinking, selective attention) and the resetting of standards, individuals high in adaptive perfectionism are more likely to perceive themselves as accomplishing standards. They can ‘rejoice in their skills’ and ‘appreciate a job well-done’, all of which ‘brings them solid satisfaction’ and serve to enhance their self-esteem (Missildine, 1963, p. 87).

In line with theory, evidence shows that adaptive perfectionism is positively correlated with self-esteem (Ashby & Rice 2002; Methikalam et al., 2015; Mobley et al., 2005; Slaney et al., 2001; Wang et al., 2007; Wang, 2012). Yet, evidence on self-esteem’s mediating role between adaptive perfectionism and psychological distress is limited and fails to paint a clear picture. Using the Almost Perfect Scale-Revised (APS-R) (Slaney., 2001), Chai et al., (2020) found support for self-esteem’s mediating role in adaptive perfectionism’s relation to depression. However, other studies that used a combination of other scales, i.e., the FMPS, fail to establish self-esteem’s mediating role (Rice et al., 1998; LaSota, 2005). Such finding could be attributed to having measured adaptive perfectionism using FMPS’s ‘personal standards’ subscale, which is believed to also measure dimensions of maladaptive perfectionism (i.e., contingent self-worth) (DiBartolo et al., 2004). Also, these two studies did not control for the effects of maladaptive perfectionism, where this dimension might have ‘contaminated’ the effects of adaptive perfectionism on psychological distress.

### Self-Compassion as a Moderator

Self-compassion involves an open, kind, and nonjudgmental attitude towards one’s suffering and inadequacies, and ‘recognizing that one’s experience is a part of common human experience’ (Neff, 2003, p. 244). Essentially, self-compassion is proposed to comprise three main components: self-kindness (vs self-criticism), mindfulness (vs over-identification), and common humanity (vs isolation). In face of failure and suffering, self-kindness entails adopting a warm and understanding attitude towards oneself instead of being self-critical. Being mindful means holding a nonjudgmental and accepting stance towards painful thoughts and feelings rather than over-identifying with them. Instead of perceiving one’s suffering as isolated from the larger and common human experience, common humanity entails seeing failures, vulnerabilities, and imperfections as normal being human (Neff, 2003). Given that the two dimensions of perfectionism differ in the way in responding to failings (self-critical vs self-accepting/flexible), the last objective of this study is to study self-compassion as a moderator, to paint a more comprehensive picture of bidimensional perfectionism and its relationship with psychological distress and self-esteem.

As maladaptive perfectionism is associated with self-criticism and ruminative tendencies when experiencing failures (Stoltz & Ashby, 2007), self-compassion should protect individuals against psychological distress when they experience failures. In particular, self-kindness could potentially buffer the harsh self-criticism when failings occur, whilst mindfulness might enable them to take a more balanced view of one’s failings and the corresponding negative internal experiences. Individuals might also feel less distressed and self-critical, recognizing that imperfections are common being human. Supporting this view, self-compassion was found to buffer its relationship with depression (Abdollahi et al., 2020; Adams et al., 2022; Ferrari et al., 2018).

As explored previously, self-esteem plays an integral role in giving rise to psychological distress amongst individuals. Lowered self-esteem linked with maladaptive perfectionism could possibly be attributed to individuals’ constant harsh self-evaluations. However, there are currently no studies that explore the moderating role of self-compassion between maladaptive perfectionism and lowered self-esteem.

It is proposed in this study that self-compassion could *buffer* the threats of maladaptive perfectionism to self-esteem in a few different ways. First, instead of reacting to mistakes with self-criticism and having their self-worth contingent on performance, self-compassion circumvents this by promoting self-kindness and unconditional acceptance (Neff, 2003). Second, being mindful also means individuals with high maladaptive perfectionism are less likely to ruminate on or over-identify with their shortcomings, seeing them as reflecting their unworthiness. Third, since maladaptive perfectionism is linked with feelings of inadequacies and the tendency to inaccurately perceive others to achieve success with minimal effort and little emotional stress (Hughes et al., 2019), common humanity could help them connect with humanity, understanding that struggles and imperfections are normal being human.

On the flip side, adaptive perfectionism is linked with positive feelings of satisfaction and motivation when striving for high standards. It is also linked with a more accepting attitude in response to failures. In particular, these are the traits that are believed to lower individuals experienced psychological distress and increase their self-esteem. Nevertheless, no studies have currently explored how self-compassion might moderate these relationships. As mentioned, inconsistent findings between adaptive perfectionism and its psychological outcomes could imply the existence of a moderating variable. For instance, previous studies might have failed to find the relationship between adaptive perfectionism and lowered psychological distress/ increased self-esteem due to not considering the level of self-compassion of individuals. This is particularly important, as theoretically, adaptive perfectionism is linked with self-acceptance in face of imperfections. During times of sufferings, self-kindness, mindfulness, and the recognition of common humanity should show a synergistic effect with adaptive perfectionism, *enhancing* its negative relationship with psychological distress and positive relationship with self-esteem.

### Research Gaps and The Present Study

Despite theories suggesting that perfectionism is bidimensional—having both a maladaptive and adaptive dimension, their corresponding psychological outcomes remain mixed. This might be due to the limited number of studies that examine maladaptive and adaptive perfectionism simultaneously, from different angles (Chai et al., 2020). Other than studying both dimensions simultaneously, the current study also aims to shed light on the two dimensions of perfectionism by exploring the *mechanism* and the *context under which* these dimensions are linked with psychological distress. This will be done by looking at self-esteem, an integral concept to perfectionism as a mediator, and self-compassion, an important way of relating to oneself as a moderator. This will be the first study to examine the constructs of bidimensional perfectionism, self-esteem, and self-compassion together. Moreover, this will also be the first study to examine self-compassion’s moderating role between bidimensional perfectionism and self-esteem, and between adaptive perfectionism and psychological distress.

Furthermore, mixed evidence on the relationship between bidimensional perfectionism and psychological distress and the mediating role of self-esteem could be attributed to a few design limitations of previous studies. They are namely: 1) the use of impure scales, 2) the failure to control the effects of maladaptive and adaptive perfectionism on one another, and 3) not taking into account the existence of other moderating variables. All of these limitations will be tackled in this study, which further contributes to the novelty of this study. It is hoped that a multi-angle exploration, together with addressing the design limitations of previous studies could enable the complex and intricate relationships between bidimensional perfectionism and psychological distress to emerge.

To address the limitation of scale use, the current study will be using the Almost Perfect Scale-Revised (APS-R). The APS-R better operationalizes the two dimensions of perfectionism for a few different reasons. First, it captures the defining features of maladaptive and adaptive perfectionism using the subscales of ‘Discrepancy’ and ‘High Standards’ respectively. This is unlike that seen in the other two major measurements, FMPS and MPS, which instead measure the causal, correlational, or the effects of being perfectionistic rather than perfectionism itself (Slaney et al., 2001). Second, it is also a ‘purer’ measure, with the subscales independently measuring the separate dimensions of maladaptive and adaptive perfectionism (Slaney et al., 2001). Third, the APS-R subscales were found to be more strongly associated with the different psychological measures, including self-esteem, depression, and worry, compared to FMPS and MPS (Slaney et al., 2001).

Lastly, given the multidimensional nature of perfectionism, the two dimensions’ relations with their psychological outcomes can only be uncovered and interpreted with a multivariate statistical approach (Stoeber & Gaudreau, 2017). Despite the two dimensions being consistently correlated, previous studies did not control for the effects on one another, where the unique effects of each dimension could not be studied closely. This also potentially contributed to the mixed literature, especially on adaptive perfectionism. Such a research gap will be addressed by statistically controlling for the effects of the other dimension, in hopes of painting a cleaner and more accurate picture of bidimensional perfectionism’s relationship with psychological distress.

### Practical Significance

Through evidencing the adaptive/maladaptive perfectionism dichotomy, this study could contribute to shifting the current mainstream psychopathological and unidimensional understanding of perfectionism. In particular, it can contribute to the refinement of psychological interventions aimed at reducing distress associated with perfectionism. Currently, perfectionism interventions are found to indiscriminately reduce both dimensions of perfectionism (Suh et al., 2019). If the two dimensions of perfectionism are found to show differential relationships with psychological distress, interventions could be refined so that they target at reducing maladaptive perfectionism whilst reinforcing and utilizing the adaptive aspects of individuals’ perfectionism.

Moreover, although these interventions, with the mainstream being Cognitive-Behavioural Therapy (CBT), were found to significantly reduce the psychological distress associated with perfectionism, the effect sizes of these studies remain moderate (Suh et al., 2019).

Moreover, post-test perfectionism levels amongst participants remain in the high range in some studies, raising questions concerning the practical significance of these interventions. These results support the view that perfectionism is a relatively stable personality trait (Cox & Enns, 2003). Contrarily, self-esteem and self-compassion are traits that can be cultivated (Ferrari et al., 2019; Niveau et al., 2021). Hence, finding their roles could enable alternative ways of intervening with distressed, perfectionistic individuals. In particular, this study can also set the stage for self-esteem/self-compassion intervention studies on perfectionism, studies that have not been conducted before.

### Hypotheses

#### Maladaptive Perfectionism

1. Maladaptive perfectionism is positively related to psychological distress.
2. Self-esteem mediates the relationship between maladaptive perfectionism and psychological distress. Higher maladaptive perfectionism is linked with lower self-esteem, which in turn is linked with higher psychological distress.
3. Self-compassion moderates the relationship between maladaptive perfectionism and self-esteem. Higher self-compassion weakens the negative relationship between maladaptive perfectionism and self-esteem.
4. Self-compassion moderates the relationship between maladaptive perfectionism and psychological distress. Higher self-compassion weakens the positive relationship between maladaptive perfectionism and psychological distress.

#### Adaptive Perfectionism

1. Adaptive perfectionism is negatively related to psychological distress.
2. Self-esteem mediates the relationship between adaptive perfectionism and psychological distress. Higher adaptive perfectionism is linked with higher self-esteem, which in turn is linked with lower psychological distress.
3. Self-compassion moderates the relationship between adaptive perfectionism and self-esteem. Higher self-compassion enhances the positive relationship between adaptive perfectionism and self-esteem.
4. Self-compassion moderates the relationship between adaptive perfectionism and psychological distress. Self-compassion enhances the negative relationship between adaptive perfectionism and psychological distress.

The hypotheses are summarized in Figures 1 and 2 below.

**Figure 1.**
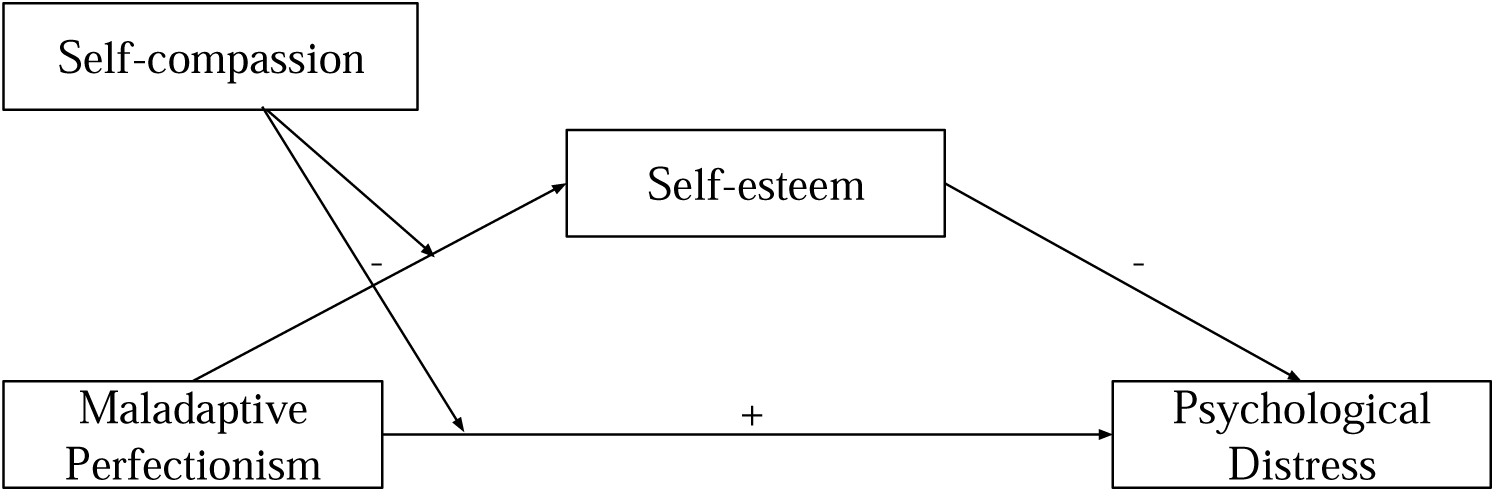
Proposed relationships among maladaptive perfectionism, psychological distress, self-esteem, and self-compassion

**Figure 2.**
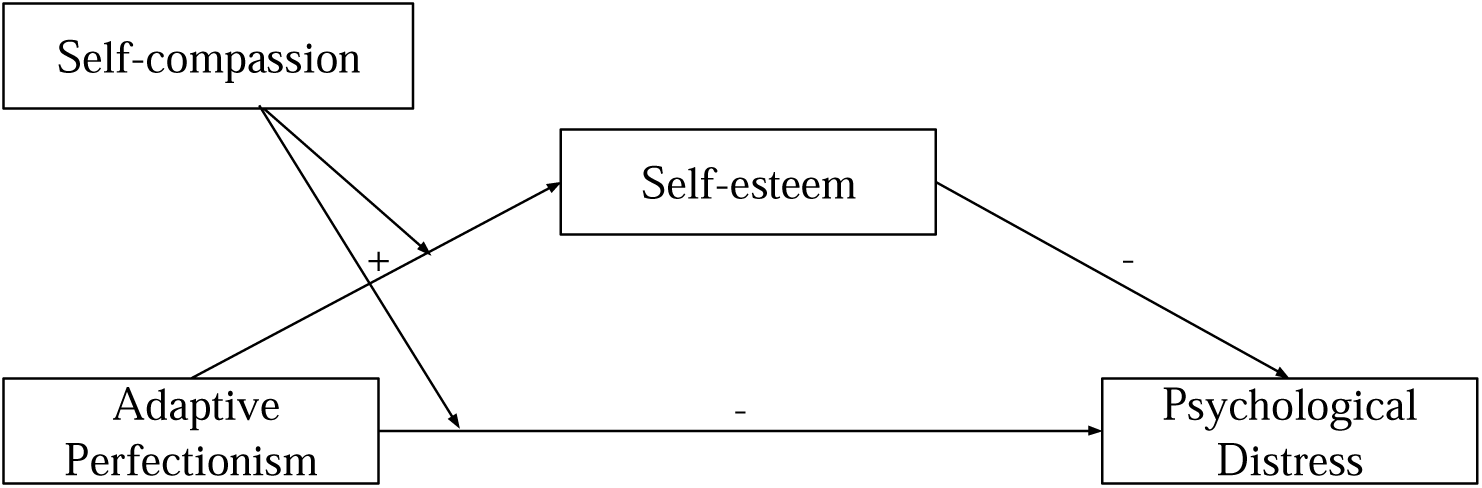
Proposed relationships among adaptive perfectionism, psychological distress, self-esteem, and self-compassion

## Methodology

### Participants

Sample size calculation for moderation analysis was conducted since the moderation effect size is smaller compared to that of mediation, Δ*R*^2^ = .021 (Ferrari et al., 2018). A priori power analysis using G*Power indicated that a minimum required sample size of 387 was needed to detect the small effect of *f* ^2^ = .02 with 80% power (α = .05).

A total of 503 participants were recruited via mass email at HKU and personal contact. As this study focuses on the general adult population, only participants above age 18 who could read English or Chinese were recruited. Participants’ mean age was 25 (SD = 7.85, range: 18-70). The majority of the participants were female (*n =* 351, 69.78%) and Asian (*n =* 435, 86.48%). Around half of the participants were studying at the time of participation (*n* = 241, 47.91%). Other demographic details are summarized in Table 1.

**Table 1.**
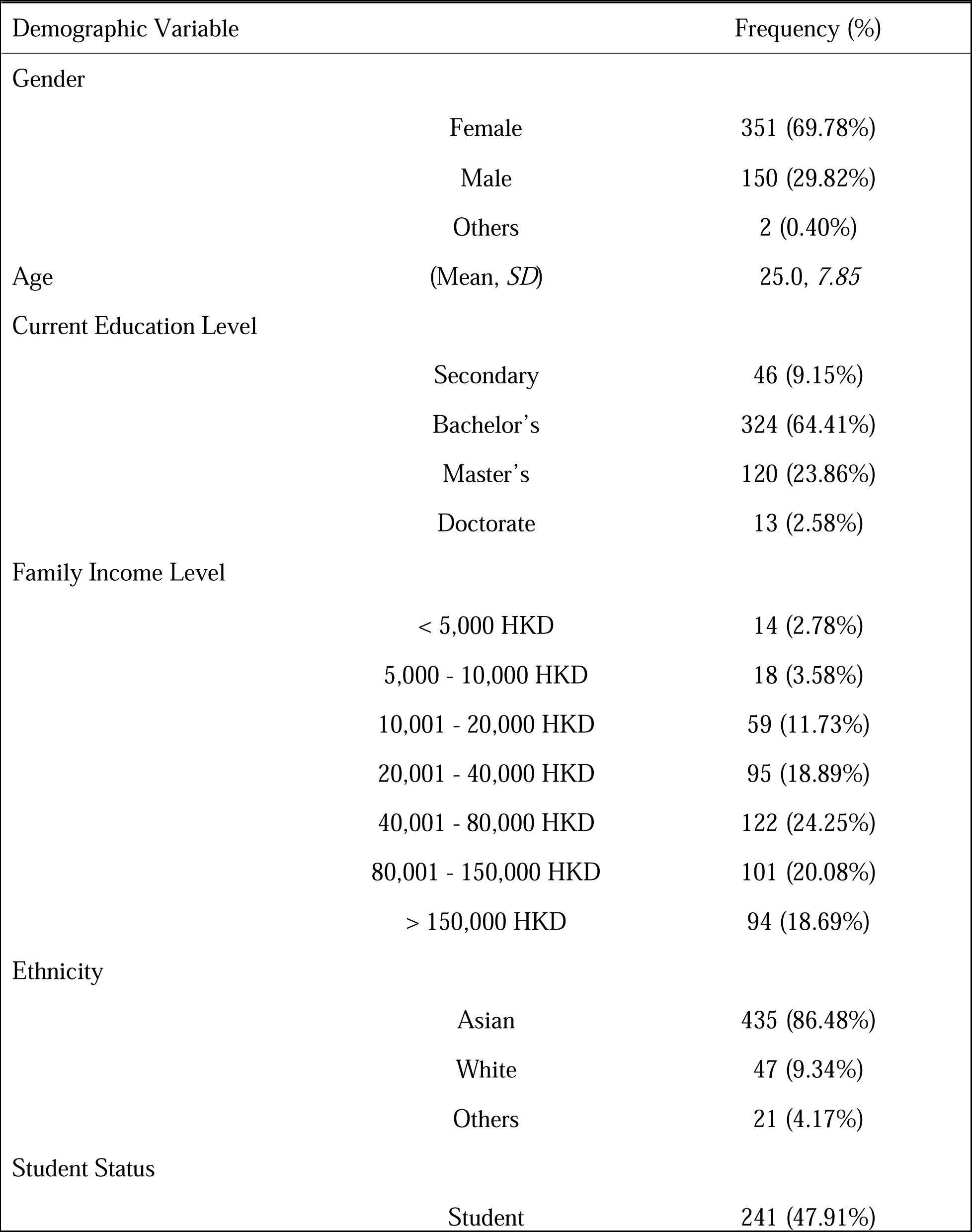
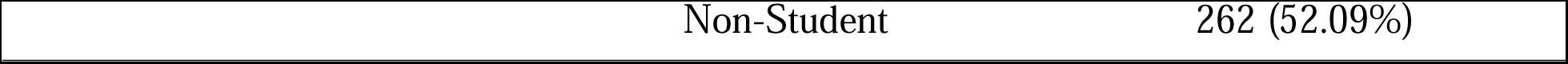
Summary of Demographic Data of Participants (N =503).

### Procedures

Ethics approval was sought prior to data collection. A Qualtrics link was disseminated via mass email at HKU and through personal contact and snowballing. Interested participants followed the link, where they were asked to fill in informed consent, basic demographics information (age, gender, ethnicity, student status, education, and family income level), and their responses to the four randomized sets of questionnaires. Participants were given a debriefing note at last to explain the nature of the study. No compensation was provided to the participants.

### Measures

#### Almost Perfect Scale-Revised (APS-R)

The 23-item APS-R (Slaney et al., 2001) was developed to be a purer, theoretically, and empirically sound measurement of bidimensional perfectionism. The subscale of ‘Discrepancy’ and ‘High Standards’ were used to measure the maladaptive and adaptive dimensions of perfectionism respectively. Participants responded to the items using a seven-point Likert rating scale ranging from 1 (*strongly disagree*) to 7 (*strongly agree*). Sample items for ‘Discrepancy’ and ‘High Standards’ include *‘I am hardly ever satisfied with my performance’* and *‘I set very high standards for myself’* respectively. The APS-R shows good internal consistencies and test-retest reliability for ‘Discrepancy’ (α = .92; .83) and ‘High Standards’ (α = .82; .72) (Grzegorek et al., 2004). The APS-R also shows good validity: ‘Discrepancy’ and ‘High Standards’ were found to positively correlate with the maladaptive and adaptive subscales of other major perfectionism subscales respectively (Slaney et al., 2001). Measures of APS-R also correlate with indicators of psychological adjustment and well-being, in directions that are consistent with theory (Slaney et al., 2001). The Chinese version of the APS-R adopted from Wang et al., (2007) also demonstrates good validity, internal consistency and test-retest reliability for ‘Discrepancy’: (α = .85 ; .71) and ‘High Standards’: (α = .76; .72) (Li et al., 2007). The Cronbach’s alphas in the current sample were .93 and .83 for ‘Discrepancy’ and ‘High Standards’ respectively.

#### Rosenberg Self-Esteem Inventory (RSE)

The RSE is comprised of 10 statements to measure a general perception of self-worth (Rosenberg, 1965). Half of the items are positively-worded and half are negatively-worded, with sample items like *‘I feel that I’m a person of worth’* and *‘I certainly feel useless at times’* respectively. Participants were scored on a four-point Likert scale ranging from 1 (*strongly agree*) to 4 (*strongly disagree*), where high scores indicate positive self-esteem. RSE shows good internal consistency (α = .81) and test-retest reliability (.85) (Schmitt & Allik, 2005; Silber & Tippett, 1965). Its validity is demonstrated by its correlation with other self-esteem measures (Goldsmith, 1986). The Chinese version the RSE adopted from Tsang (1997), amended by (Leung, 2008) also demonstrates good validity and reliability (α = 0.76). The Cronbach’s alpha was .86 in the current sample.

#### Self-Compassion Scale (SCS)

The 26-item SCS (Neff, 2003) was used to measure self-compassion. It comprises 6 subscales: ‘Self-Kindness’, ‘Self-Judgment’, ‘Common Humanity’, ‘Isolation’, ‘Mindfulness’, and ‘Over-identification’. Participants responded on a 5-point Likert scale ranging from 1 (*almost never*) to 5 (*always*) in terms of how often they behave in the stated manner. A high score demonstrates high self-compassion. A sample item for ‘Self-Kindness’ includes *‘I try to be loving towards myself when I’m feeling emotional pain’*, that of ‘Common Humanity’ includes *‘I try to see my failings as part of the human condition’*, and that of ‘Mindfulness’ includes *‘When something upsets me I try to keep my emotions in balance’*. Total SCS scores evidenced good internal reliability (α = .92) and test-retest reliability (.93) (Neff, 2003). SCS was also consistently found to positively link with psychological well-being, suggesting its validity (Zessin et al., 2015). The Chinese version of the SCS also demonstrates good validity, internal consistency (α = .84), and test-retest reliability (.89) (Chen & Zhou, 2011). The Cronbach’s alpha was .92 in the current sample.

#### Kessler Psychological Distress Scale (K10)

Psychological distress was measured using the 10-item K10 (Kessler, 1996). K10 measures the frequency of depressive and anxiety symptoms in the past four weeks, where participants responded on 5-point Likert scale, ranging from 1 (*none of the time*) to 5 (*all the time*). Sample items for measuring depressive and anxiety symptoms include *‘How often did you feel hopeless?’* and *‘How often did you feel nervous*?*’* respectively. K10 demonstrates good validity, internal reliability (α = .92), and test-retest reliability (.89) (Kong et al., 2021; Merson et al., 2021). The Chinese version of the K10 also demonstrates high validity (α = .80) and test-retest reliability (.70) (Zhou et al., 2008). The Cronbach’s alpha was .92 in the current sample.

### Statistical Analysis

IBM SPSS statistics 28 was used to obtain descriptive statistics, intercorrelations among variables, and to conduct mediation and moderation analyses. Hayes (2013)’s models 4 and 8 in the PROCESS template were used for mediation and moderation analyses respectively following the bias-corrected bootstrapping method. 5000 samples were randomly sampled from the original data to estimate bias-corrected standard errors and the 95% confidence intervals. A 95% confidence interval (CI) without zero indicates a statistically significant result at a level of α = 0.05. Additionally, gender, education, and family income levels were translated into dummy variables and were added as covariates in these two models.

## Results

### Sample Characteristics

The sample size was 503, with 351 females and 150 males (2 identified as ‘others’) aged between 18 to 70 years (*M* = 25.0, *SD =* 7.85). Table 1 also shows the other background characteristics including ethnicity, education, and family income levels that were then tested as potential confounding variables.

Independent sample t-tests showed that there were no significant gender differences in any of the study variables. Student status differences were only found for self-esteem, with those who were students (*M =* 27.55, *SD =* 5.16) demonstrating significantly lower self-esteem than those who were not (*M* = 27.91, *SD* = 4.56), *t*(501) = -.84, *p* = .01.

One-way ANOVA revealed there were significant differences between education levels for adaptive perfectionism, *F*(3, 499) = 4.15, *p =* .01, maladaptive perfectionism, *F*(3, 499) = 3.81, *p* = .01, self-esteem, *F*(3, 499) = 5.94, p < .001, psychological distress *F*(3, 499) = 3.65, *p =* .013 and self-compassion, *F*(3, 499) = 3.07, *p = .*027. There were also significant differences between family income levels for maladaptive perfectionism, *F*(6, 496) = 2.30, *p* = .03, and psychological distress *F*(6, 496) = 2.34 *p* = .03. Hence, education and family income levels were added as covariates in subsequent mediation and moderated analyses.

### Preliminary Correlation Analysis

Table 2 shows the means, standard deviations and zero-order correlations of the main study variables and age. Without controlling for adaptive perfectionism and other demographic variables, maladaptive perfectionism had significant positive correlations with adaptive perfectionism (*r* = .31, *p <* .01), psychological distress (*r* = .54, *p <* .01), and significant negative correlations with self-esteem (*r* = -.65, *p <* .01), self-compassion (*r* = -.61, *p <* .01), and age (*r* = -.22, *p <* .01).

**Table 2.** Summary of Means, Standard Deviations, and Zero-Order Correlations of the Main Variables and Age.

|  | *M* | *SD* | 1 | 2 | 3 | 4 | 5 | 6 |
| --- | --- | --- | --- | --- | --- | --- | --- | --- |
| 1. Maladaptive Perfectionism | 50.53 | 14.29 | - | .31** | -.65** | -.61** | .54** | -.22** |
| 2. Adaptive Perfectionism | 37.98 | 6.73 |  | - | .05 | -.21** | .14** | -.23** |
| 3. Self-Esteem | 27.75 | 4.86 |  |  | - | .64** | -.56** | .12** |
| 4. Self-Compassion | 3.01 | .62 |  |  |  | - | -.55** | .21** |
| 5. Psychological Distress | 24.85 | 7.25 |  |  |  |  | - | -.14** |
| 6. Age | 25.04 | 7.85 |  |  |  |  |  | - |

\*\* $p < .01$ (2-tailed)

Without controlling for maladaptive perfectionism and other demographic variables, adaptive perfectionism had a significant positive correlation with psychological distress (*r* = .14, *p <* .01), and significant negative correlations with self-compassion (*r* = -.21, *p <* .01) and age (*r* = -.23, *p <* .01). Since age was significantly correlated with all the study variables, it was also added as a covariate for further analyses. Moreover, since maladaptive perfectionism and self-compassion were significantly correlated, to reduce multicollinearity, mean-centered moderation analyses were conducted to test hypotheses 3 and 4.

### Mediation Analysis

Hayes’ (2013) model 4 in the PROCESS macro was employed to test for hypotheses 1, 2, 5, and 6.

To test hypotheses 1 and 2, maladaptive perfectionism was entered as the independent variable, psychological distress as the dependent variable, and self-esteem as the mediator.

Adaptive perfectionism, age, education, and family income levels were added as covariates. Results are summarized in Table 3 and the path diagram can be found in Figure 3.

**Figure 3.**
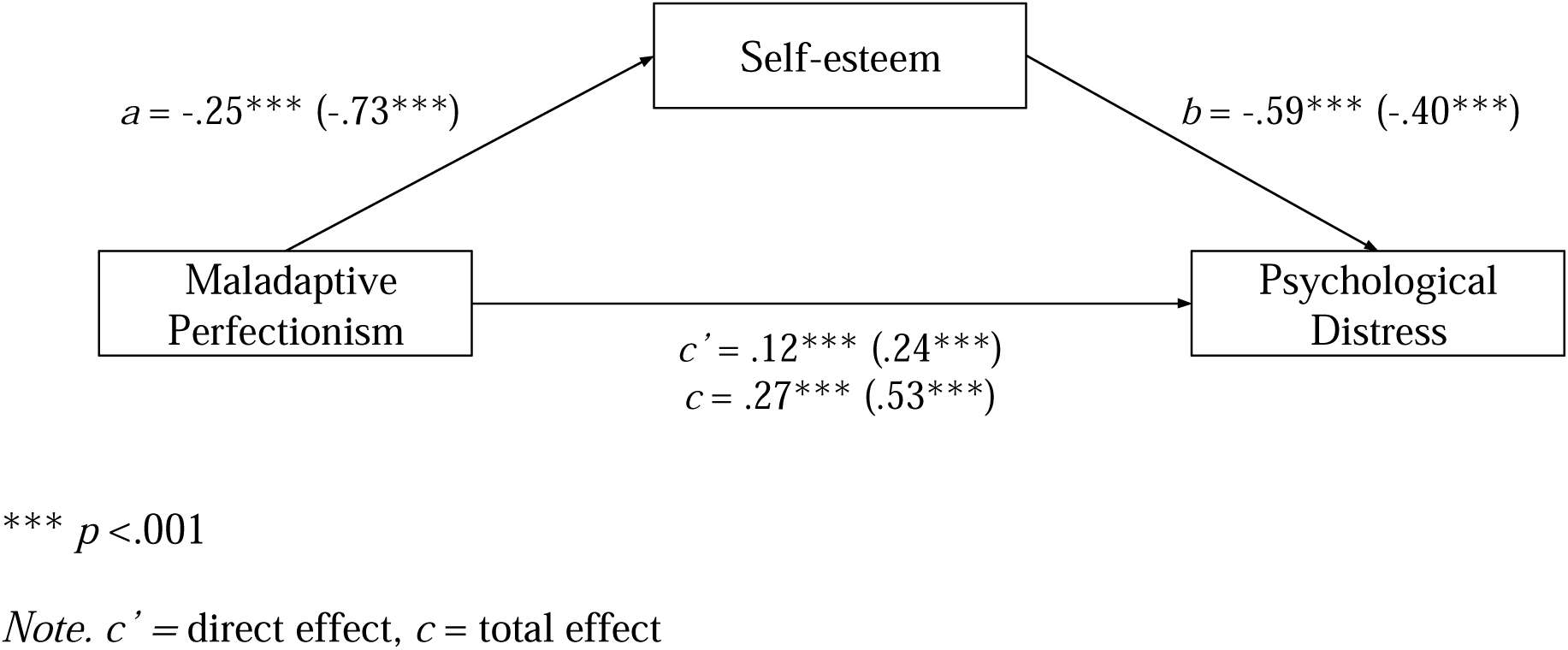
A path diagram of the mediation model in which maladaptive perfectionism is associated with psychological distress through self-esteem. Note. Numerical values in parentheses are standardized path coefficients. Those not in parentheses are unstandardized path coefficients.

**Table 3.** Total, Direct, Indirect Effects and 95% CI from Mediation Analysis Self-Esteem as a Mediator Between Maladaptive Perfectionism and Psychological Distress.

| | $b$ | $\beta$ | $SE$ | $p$ | 95% CI |
| --- | --- | --- | --- | --- | --- |
| Total Effect | .27 | .53 | .02 | < .001 | **[.23, .31]** |
| Direct Effect | .12 | .24 | .03 | < .001 | **[.07, .18]** |
| Indirect Effect | .15 | .29 | .02 | N/A | **[.11, .19]** |

*Note.* Bolded means significant results.

#### Hypothesis 1: Maladaptive perfectionism is positively related to psychological distress

The total effect model showed that maladaptive perfectionism significantly positively predicted psychological distress, *b* = .27, β = .53, 95% CI [.23, .31], *p <* .001. Hypothesis 1 was supported.

#### Hypothesis 2: Self-esteem mediates the relationship between maladaptive perfectionism and psychological distress. Higher maladaptive perfectionism is linked with lower self-esteem, which in turn is linked with higher psychological distress

Results of the bias-corrected bootstrapped analyses found that maladaptive perfectionism had a significant indirect effect on distress via self-esteem, *b =* .15, β = .29, 95% CI [.11, .19]. In particular, participants with higher levels of maladaptive perfectionism showed lower self-esteem, *b* = -.25, β = -.73, 95% CI [-.27, -.23], *p <* .001, and those who showed lower self-esteem reported greater distress, *b* = -.59, β = -.40, 95% CI [-.74, -.45], *p <* .001. Hypothesis 2 was supported.

Another mediation analysis was conducted to test hypotheses 5 and 6. Adaptive perfectionism was entered as the independent variable, psychological distress as the dependent variable, and self-esteem as the mediator. Maladaptive perfectionism, age, education, and family income levels were added as covariates. Results are summarized in Table 4 and Figure 4.

**Figure 4.**
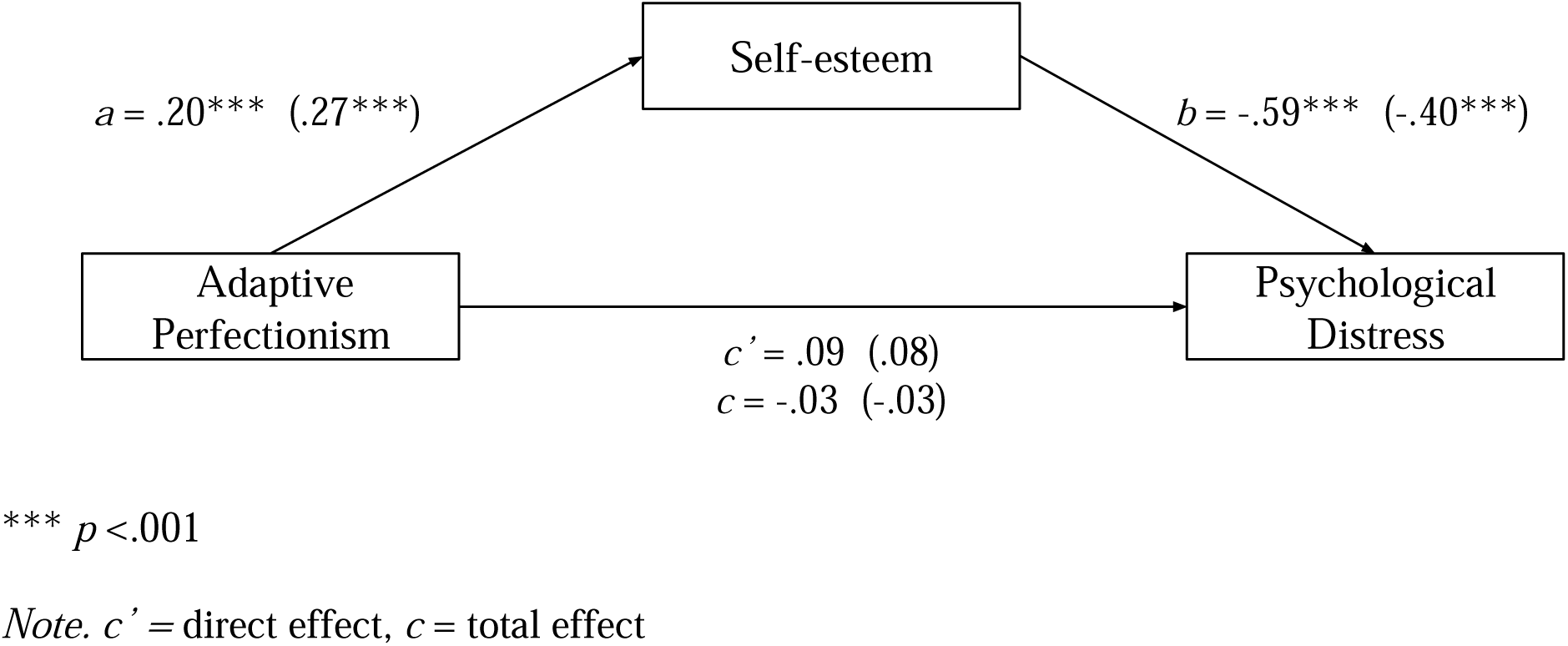
A path diagram of the mediation model in which adaptive perfectionism is associated with psychological distress through self-esteem. Note. Numerical values in parentheses are standardized path coefficients. Those not in parentheses are unstandardized path coefficients.

**Table 4.** Total, Direct, Indirect Effects and 95% CI from Mediation Analysis Self-Esteem as a Mediator Between Adaptive Perfectionism and Psychological Distress.

| | *b* | $\beta$ | *SE* | *p* | 95% CI |
| --- | --- | --- | --- | --- | --- |
| Total Effect | -.03 | -.03 | .04 | .50 | [-.12, .06] |
| Direct Effect | .09 | .08 | .04 | .05 | [-.002, .17] |
| Indirect Effect | -.12 | -.11 | .02 | N/A | **[-.16, -.08]** |

Note. Bolded means significant results.

#### Hypothesis 5: Adaptive perfectionism is negatively related to psychological distress

The total effect model showed that adaptive perfectionism was unrelated to distress, *b* = -.03, β = -.03, 95% CI [-.12, .06], *p* = .50. Hypothesis 4 was not supported.

#### Hypothesis 6: Self-esteem mediates the relationship between adaptive perfectionism and psychological distress. Higher adaptive perfectionism is linked with higher self-esteem, which in turn is linked with lower psychological distress

Results of the bias-corrected bootstrapped analyses found that adaptive perfectionism had a significant indirect effect on distress via self-esteem, *b* = -.12, β = -.11, 95% CI [-.16, -.08]. In particular, participants with higher levels of adaptive perfectionism showed higher self-esteem, *b* = .20, β = .27, 95% CI [.15, .25], *p <* .001, and those who showed higher self-esteem experienced lower psychological distress, *b* = -.59, β = -.40, 95% CI [-.74, -.45], *p <*. 001. Hypothesis 5 was supported.

### Moderation Analysis

#### Hypothesis 3: Self-compassion moderates the relationship between maladaptive perfectionism and self-esteem. Higher self-compassion weakens the negative relationship between maladaptive perfectionism and self-esteem

The overall model was significant, *R* = .78, *F*(15, 487) = 50.33, *p* < .001. The inclusion of the product term (maladaptive perfectionism × self-compassion) into the model results in a significant increase in variance accounted for by the predictor variables, Δ*R*^2^ = .0038, *F* (1, 488) = 4.72, *p* = .030. This indicates that self-compassion significantly moderated the relationship between maladaptive perfectionism and self-esteem. To probe the significant interaction, simple slopes analyses were conducted, and the results are visualized in Figure 5. The negative relationship between maladaptive perfectionism and psychological distress were significant at one SD below the mean self-compassion score, *b* = -.19, *t* = -12.09, 95% CI [-.22, -.16], *p* < .001, at the mean self-compassion score, *b =* -.16, *t* = -12.67, *p < .*001, 95% CI [-.19, -.14], and at one SD above the mean self-compassion score, *b* = -15, *t* = -9.10, *p < .*001, 95% CI [-.18, -.11].

**Figure 5.**
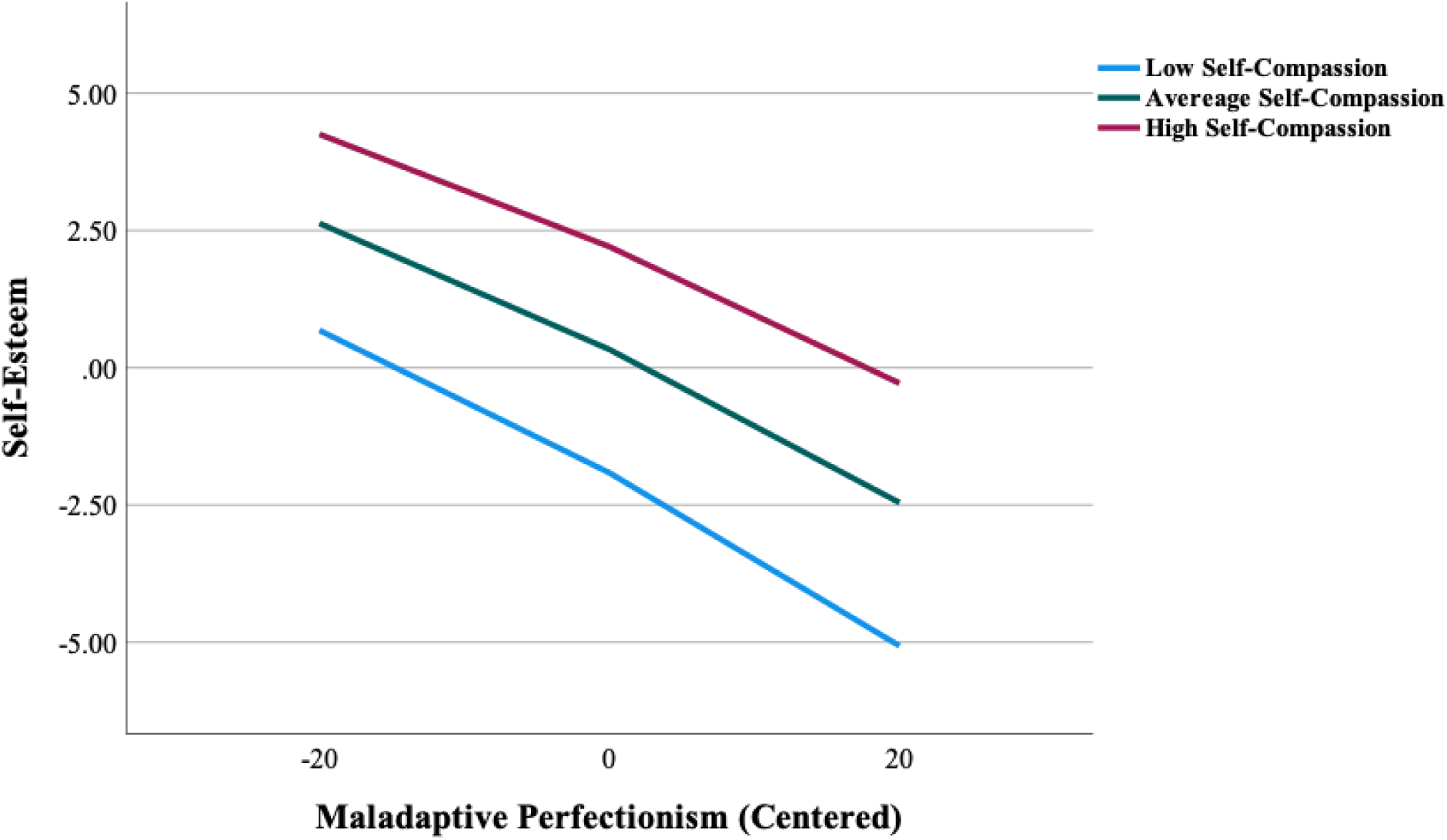
The moderating effect of self-compassion on maladaptive perfectionism and self-esteem.

**Table 5.**
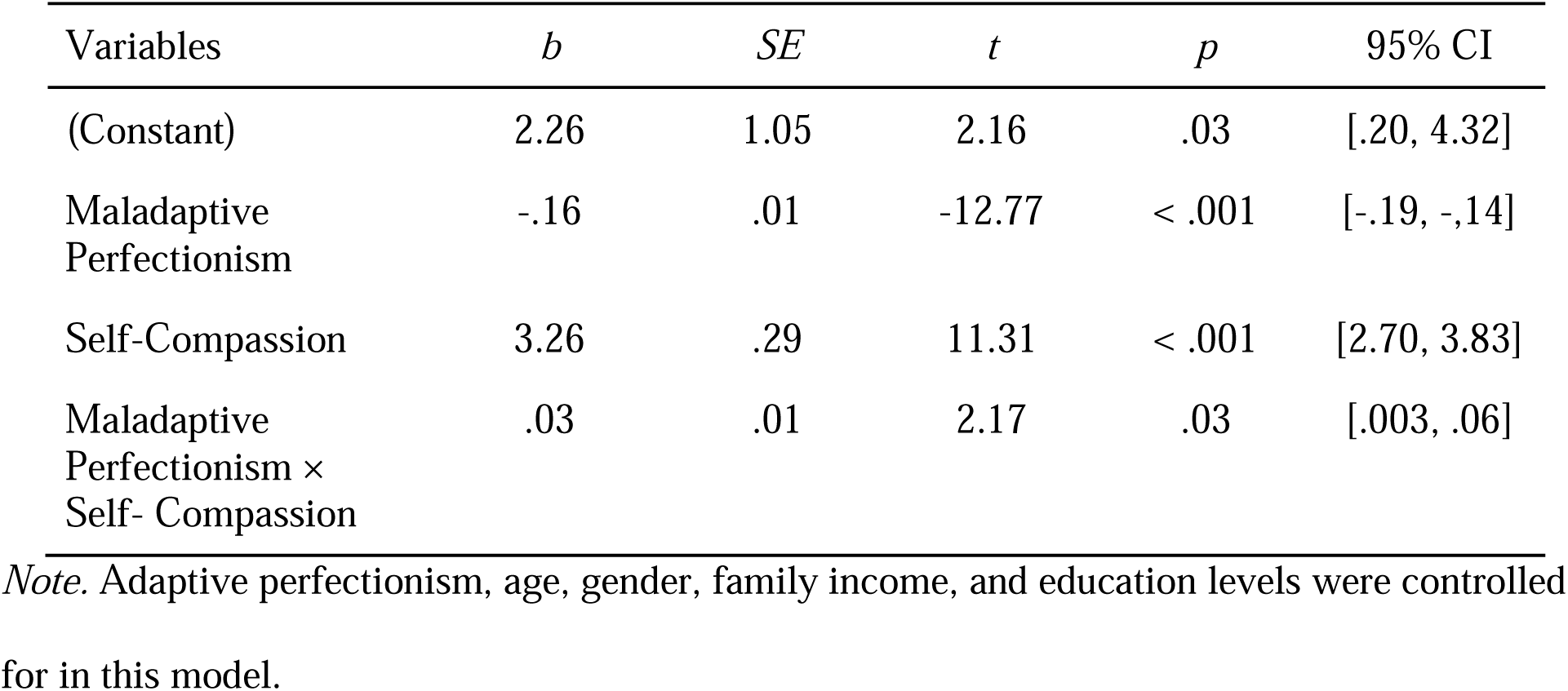
Moderation Analysis for Self-Compassion in the Relationship Between Maladaptive Perfectionism and Self-Esteem.

However, the higher the self-compassion, the weaker relationship between maladaptive perfectionism and self-esteem was found, as reflected by a less steep slope in the graph. Hypothesis 3 was supported.

#### Hypothesis 4: Self-compassion moderates the relationship between maladaptive perfectionism and psychological distress. Higher self-compassion weakens the positive relationship between maladaptive perfectionism and psychological distress

The overall model was significant, *R* = .65, *F*(15, 487) = 23.15, *p* < .001. However, the inclusion of the product term (maladaptive perfectionism × self-compassion) into the model did not result in a significant increase in variance accounted for by the predictor variables, Δ*R*^2^ = .0006, *F* (1, 487) = .50, *p* = .48. Hypothesis 4 was not supported.

**Table 6.** Moderation Analysis for Self-Compassion in the Relationship between Maladaptive Perfectionism and Psychological Distress.

| Variables | *b* | *SE* | *t* | *p* | 95% CI |
| --- | --- | --- | --- | --- | --- |
| (Constant) | -.66 | 1.92 | -.35 | .73 | [- 4.43, 3.10] |
| Maladaptive Perfectionism | .09 | .03 | 3.39 | < .001 | [.04, .15] |
| Self-Compassion | -3.01 | .59 | -5.09 | < .001 | [-54.17, -1.84] |
| Maladaptive Perfectionism × Self-Compassion | -.02 | .03 | -.72 | .47 | [-.07, .03] |

*Note.* Adaptive perfectionism, age, gender, family income, and education levels were controlled for in this model.

#### Hypothesis 7: Self-compassion moderates the relationship between adaptive perfectionism and self-esteem. Higher self-compassion enhances the positive relationship between adaptive perfectionism and self-esteem

The overall model was significant, *R* = .64, *F*(15, 487) = 23.09, *p* < .001. However, the inclusion of the product term (adaptive perfectionism × self-compassion) into the model did not result in a significant increase in variance accounted for by the predictor variables, Δ*R*^2^ = .002, *F* (1, 487) = .18, *p* = .89. Hypothesis 7 was not supported.

**Table 7.** Moderation Analysis for Self-Compassion in the Relationship Between Adaptive Perfectionism and Self-Esteem.

| Variables | *b* | *SE* | *t* | *p* | 95% CI |
| --- | --- | --- | --- | --- | --- |
| (Constant) | 14.50 | 4.31 | 3.37 | < .001 | [6.04, 22.96] |
| Adaptive Perfectionism | .38 | .10 | 3.63 | < .001 | [.17, .59] |
| Self-Compassion | 5.53 | 1.33 | 4.15 | < .001 | [2.91, 8.14] |
| Adaptive Perfectionism × Self- Compassion | -.06 | .03 | -1.80 | .074 | [-.13, .006] |

*Note.* Maladaptive perfectionism, age, gender, family income, and education levels were controlled for in this model.

#### Hypothesis 8: Self-compassion moderates the relationship between adaptive perfectionism and psychological distress. Self-compassion enhances the negative relationship between adaptive perfectionism and psychological distress

The overall model was significant, *R* = .78, *F*(14, 488) = 53.76, *p* < .001. However, the inclusion of the product term (maladaptive perfectionism × self-compassion) into the model did not result in a significant increase in variance accounted for by the predictor variables, Δ*R*^2^ = .00002, *F* (1, 488) = 3.21, *p* = .07. Hypothesis 8 was not supported.

**Table 8.** Moderation analysis for Self-Compassion in the Relationship Between Maladaptive Perfectionism and Psychological Distress.

| Variables | *b* | *SE* | *t* | *p* | 95% CI |
| --- | --- | --- | --- | --- | --- |
| (Constant) | 38.79 | 7.93 | 4.89 | < .001 | [23.20, 54.37] |
| Adaptive Perfectionism | .02 | .19 | .12 | .91 | [-.36, .40] |
| Self-Compassion | -3.26 | 2.47 | -1.32 | .19 | [-9.16, 1.60] |
| Adaptive Perfectionism × Self- Compassion | .008 | .06 | .14 | .90 | [-.11, .12] |

*Note.* Maladaptive perfectionism, age, gender, family income, and education levels were controlled for in this model.

## Discussion

Although there is a consensus that perfectionism has both adaptive and maladaptive dimensions, empirical evidence on their relationships with psychological outcomes remains mixed. The objective of this study is to provide an in-depth examination of the two dimensions’ relationships with psychological distress. This was done by (1) studying the two dimensions of perfectionism simultaneously with regards to their relationship to psychological distress, (2) exploring the mediating role of self-esteem, (3) exploring the role of self-compassion on their relationships with self-esteem and psychological distress, and (4) addressing the limitations of previous research: using the APS-R and controlling for the effects of the other dimension.

Results showed that maladaptive perfectionism was positively linked with psychological distress, whilst adaptive perfectionism was not related to psychological distress. Self-esteem was also found to mediate both dimensions of perfectionism with psychological distress, in the expected direction. Self-compassion only moderated the relationship between maladaptive perfectionism and self-esteem but not that between maladaptive perfectionism and psychological distress. It also did not moderate the relationships between adaptive perfectionism with psychological distress and self-esteem. Major findings are discussed below.

### Maladaptive Perfectionism

#### Maladaptive Perfectionism - Psychological Distress

As hypothesized, results showed that maladaptive perfectionism positively predicted psychological distress. This adds to the existing literature and reinforces our current understanding that maladaptive perfectionism is indeed maladaptive. Previously, most studies (e.g., Dunkley et al., 2000., Einstein et al., 2001) that also found such a result operationalized maladaptive perfectionism using the FMPS and MPS, suggesting that maladaptive perfectionism might be linked to psychological distress due to individuals’ excess doubts over action, concerns over mistakes, and the perception that others have unrealistically high standards on them. By using the APS-R, Kong et al., (2021), Wang et al., (2009) and the current study further suggest that it is also the perceived discrepancy between one’s high expectations and perceived performance, the apprehension about meeting high standards and dissatisfaction with oneself that might contribute to symptoms of depression and anxiety. Additionally, the current study extended these two studies by adopting a more statistically rigorous method: controlling for the effects of adaptive perfectionism. Controlling for the effects of adaptive perfectionism while examining the relationship between maladaptive perfectionism and psychological distress allows the unique contributions of maladaptive perfectionism on psychological distress to be dissected.

#### Self-Esteem as a Mediator Between Maladaptive Perfectionism and Psychological Distress

Self-esteem was found to mediate the relationship between maladaptive perfectionism and psychological distress. More specifically, those who have high maladaptive perfectionism are more likely to experience lowered self-esteem, where this in turn contributes to greater psychological distress. This is as expected, as theoretically, maladaptive perfectionism is linked with inflexible thinking styles and the tendency to perceive slight discrepancies from high standards as evidence that one has failed (Flett & Hewitt, 2002). When the constant perceived discrepancy is internalized, it leads to constant feelings of dissatisfaction and unworthiness, as reflected by lowered self-esteem (Halgin & Leahy, 1989). The current result echoes that of previous studies, showing that maladaptive perfectionism is linked with lowered self-esteem (Ashby & Rice, 2002; Rice et al., 1998). Moreover, according to the vulnerability model, supported by empirical longitudinal evidence (Sowislo & Orth, 2013), low self-esteem put individuals at risk for both depression and anxiety during stressful life events (Mu et al., 2019). One explanation is that low self-esteem gives rise to negative beliefs about the self, leading to the development of depression and anxiety (Beck, 1967).

Previously, mediational studies of self-esteem between maladaptive perfectionism and psychological distress have yielded mixed results. It was initially proposed that this could be due to the use of varying and impure scales. Whilst Chai et al. (2020), Zhang & Cai (2012), Moroz & Dunkley (2015) and Preusser et al.’s (1994) studies that employed the purer APS-R scale found such a mediating role of self-esteem in both Eastern and European community samples, Rice et al. (1998) and Park et al.’s, (2010) studies that used APS (not revised) and MPS did not. APS-R was employed in the current study, and as expected, yielded positive results. This suggests that APS-R might also be a purer scale in measuring maladaptive perfectionism, instead of just being a purer scale for adaptive perfectionism. The APS-R could better operationalize maladaptive perfectionism and capture its related psychological outcomes due to its measurement of one’s perceived discrepancy, a theoretically integral characteristic of maladaptive perfectionism (Horney, 1951). Additionally, by controlling for the effects of adaptive perfectionism, this study further shows the mediating role of self-esteem exists between the independent trait of maladaptive perfectionism and psychological distress, which is a strength of this study.

#### Self-Compassion as a Moderator Between Maladaptive Perfectionism and Self-Esteem

Although self-compassion should theoretically buffer the negative link between maladaptive perfectionism and self-esteem, no study has tested this supposition previously. The current study suggests that such moderating effect exists, reflecting that although maladaptive perfectionism might lead to lowered self-esteem, such a relationship is weakened when individuals also have higher self-compassion. Self-compassion entails being kind, balanced, and understanding towards oneself during suffering (Neff, 2009). Self-compassion might be able to attenuate the negative self-evaluations and ruminative tendencies linked with maladaptive perfectionism, tendencies that could give rise to feelings of unworthiness when personal failings occur.

Similar effects of self-compassion were also found in Leary et al., (2007)’s research. In their study 2 and 3, self-compassion was found to buffer people against negative self-feelings when imagining distressing social events and after receiving ambivalent feedback. These authors suggested that self-compassion might help individuals accept undesirable aspects of their character more readily instead of obsessing over them. Similarly, the same mechanism of self-compassion might also help promote self-acceptance and attenuate obsession over imperfections’ implications on one’s self-worth and hence self-esteem. Moreover, in their study 4, these authors also found that people who were more self-compassionate had a more accurate perception of their video-taped performances. Applying it to the context of perfectionism, the more realistic and balanced evaluation of oneself linked with higher self-compassion might help buffer maladaptive perfectionists’ tendencies to undervalue and be biased of their performances, leading to an overall sense of unworthiness.

#### Self-Compassion as a Moderator Between Maladaptive Perfectionism and Psychological Distress

Contrary to initial prediction, self-compassion did not buffer the positive direct relationship between maladaptive perfectionism and psychological distress. This is unexpected, as self-compassion should buffer the psychological distress following the self-criticism, dissatisfaction, and apprehension linked with maladaptive perfectionism. However, this could be due to using a composite scale that measures depressive and anxiety symptoms globally as an indication of overall psychological distress. Despite being highly related, depression and anxiety are conceptually distinct and can be empirically distinguished (Sowislo et al., 2013). The tripartite model of anxiety and depression (Clark & Watson, 1991) posits that although depression and anxiety share the feature of high negative affectivity (tendency to experience nonspecific psychological distress), heightened automatic arousal is specific to anxiety, whereby low positive affectivity (e.g., loneliness, sadness) characterizes depression. Looking more closely at the construct of self-compassion, it entails having a kind and non-judgmental attitude towards ourselves when we suffer, fail, or feel inadequate (Neff, 2009). The scale expresses ‘sufferings’ in terms like *‘when I’m feeling down’, ‘when something upsets me’,* and *‘people are probably happier than I am’* etc., which are ‘sufferings’ that relate more to the feelings of sadness and low mood (i.e., low positive affectivity) as seen in depression, rather than the worries and apprehension (i.e., physiological hyperarousal) as seen in anxiety. Hence, the buffering effect of self-compassion might be stronger on depression than on anxiety, where employing a composite measure in this study might have diluted such an effect. In fact, existing literature supports this interpretation. The two studies that looked at the relationship between maladaptive perfectionism and depression found the protective effect of self-compassion (Abdollahi et al., 2020; Ferrari et al., 2018), yet the one that looked at perfectionism and psychological distress (depression and anxiety symptoms combined) did not (Ong et al., 2021). With adaptive perfectionism controlled for, the current study further informs future research that self-compassion might have unique moderating effects on maladaptive perfectionism’s relationships with depression and anxiety respectively.

Moreover, it is also possible that self-compassion is unable to buffer some dimensions of maladaptive perfectionism. Although self-compassion protected against lowered self-esteem, from the mediation analysis, the direct effect between maladaptive perfectionism and psychological distress was significant, suggesting the existence of other mediators of the relationship between maladaptive perfectionism and psychological distress. Perhaps self-compassion was unable to buffer some of these other mediators, e.g., low frustration tolerance (Hamachek, 1978) and social comparison (Saadat et a., 2017), hence failing to buffer the direct effect between maladaptive perfectionism and psychological distress.

### Adaptive Perfectionism

#### Adaptive Perfectionism – Psychological Distress

Adaptive perfectionism was hypothesized to be negatively linked to psychological distress. Addressing the limitations of previous studies, namely using APS-R and controlling for the effects of maladaptive perfectionism, such an effect was not observed in the current study.

This failed to replicate the results of Abdollahi et al., (2020), where a positive link between adaptive perfectionism and depression was found. However, this could be due to Abdollahi et al., (2020) using a sample of depressed patient, whilst the current findings are based on a non-clinical sample.

The current results echo the majority of the studies that also found a lack of relationship between adaptive perfectionism and negative psychological outcomes (e.g., Bieling et al., 2004; Frost et al., 1993). In particular, despite addressing the limitation of these studies by using the purer measurement of APS-R and controlling for maladaptive perfectionism, adaptive perfectionism still does not seem to protect individuals against depressive and anxiety symptoms. However, whilst traits like being motivated to succeed and feeling satisfied with one’s high achievements are deemed positive, they might not necessarily reduce the amount of psychological distress individuals face. Rather, these traits might be more related to positive psychological outcomes, for example, greater life satisfaction, presence of meaning, self-efficacy, positive affect (Shih, 2011; Suh et al., 2017), and higher self-esteem in the current study. Perhaps the ‘adaptiveness’ of adaptive perfectionism lies in its ability to give rise to positive psychological outcomes, rather than lower psychological distress. In terms of negative psychological outcomes in this study, the differential relationship between the two dimensions of perfectionism and psychological distress is still in keeping with the bidimensional view of perfectionism (Gäde et al., 2017).

It is important to note that, before controlling for maladaptive perfectionism, zero-order correlation showed that adaptive perfectionism had a significant positive relationship with psychological distress. This might explain why some studies found positive links between adaptive perfectionism and depression/anxiety (e.g., Bieling et al., 2004; Dunkley et al., 2003; Hill et al., 2004). Previous arguments that adaptive perfectionism might also be negative should be revisited with the awareness of the ‘contaminating’ effects of maladaptive perfectionism (e.g., self-criticism, and dissatisfaction). This is especially important as the two dimensions are consistently found to be positively correlated (Stoeber & Otto, 2006), which was also the case in this study.

#### Self-Esteem as a Mediator Between Adaptive Perfectionism and Psychological Distress

Self-esteem was found to mediate the relationship between adaptive perfectionism and psychological distress. Adaptive perfectionism is linked with higher self-esteem, probably due to individuals’ tendency to strive for high standards, whilst being motivated to improve and show the ability to be satisfied upon achievement. Existing literature supports this view, finding that high academic achievements (Aryana, 2010), being intrinsically motivated to learn (Murphy & Roopchand, 2003), and having a growth mindset (KyoungHwang & Lee, 2018) all predict higher self-esteem. A mediating role of self-esteem, then, also means that such increased self-esteem could protect individuals against psychological distress. In response to stress (e.g., underachievement), individuals with higher self-esteem might resort to more adaptive coping (Lane et al., 2002) and exhibit less self-criticism (Gittins & Hunt, 2020), protecting one against psychological distress. Adaptive perfectionism was in fact found to positively predict challenge appraisal and active coping (Stoeber & Rennert, 2008), and less self-criticism (Lo & Abbott, 2019).

Whilst previous studies that used the FMPS scale failed to find such a mediating role, this study, together with Chai et al., (2020)’s study that employed the APS-R found the mediating role of self-esteem. This suggests the previously argued importance of employing a purer measurement of adaptive perfectionism (APS-R). Additionally, since the effects of maladaptive perfectionism were controlled for in this study, stronger conclusions regarding self-esteem’s mediating role between the trait of adaptive perfectionism and psychological distress could be made.

Although there was a significant indirect effect of self-esteem, the total effect between adaptive perfectionism and psychological distress was insignificant. This suggests that there exist other pathways through which adaptive perfectionism was linked with higher psychological distress, canceling out the positive effects of increased self-esteem. One of the possible pathways could be the pressure to maintain high standards. The APS-R ‘High Standards’ subscale measures adaptive perfectionism in terms of striving for high standards, e.g., with questions like *‘I have a strong need to strive for excellence’*, *‘I expect the best from myself’*. Meanwhile, the ‘Discrepancy’ subscale measures maladaptive perfectionism in terms of the perceived inability to achieve one’s standards, or the dissatisfaction that follows, e.g., with questions like *‘I rarely live up to my high standards’*, *‘I am hardly ever satisfied with my performance’*. Hence, despite controlling for maladaptive perfectionism, such measurement can still capture distressed ‘high achievers’ that *are* able to achieve their high standards, *do* feel satisfied with attaining their goals, yet are under the constant pressure to maintain these high standards. For these individuals, the striving for high standards might then create greater psychological distress, especially feelings of anxiety. In fact, it has been argued that individuals high in self-esteem are also vulnerable individuals, since they are characterized by conditional self-acceptance, excessive focus on evaluation, and social comparisons with others (Ellis, 1995). Although high self-esteem statistically, *in isolation*, might be linked with reduced psychological distress, these individuals might also be preoccupied with obtaining the approval and avoiding disapproval from other people, suffering from the psychological distress of needing to maintain high standards. This is seen in-real life case studies in Flett and Hewitt (2002)’s research.

#### Self-Compassion as a Moderator between Adaptive Perfectionism and Self-Esteem

Self-compassion did not moderate the relationship between adaptive perfectionism and self-esteem. One possible reason is that self-compassion refers to treating oneself kindly during times of sufferings, e.g., showing the ability to buffer self-criticism linked with maladaptive perfectionism. However, although adaptive perfectionism is linked with less self-criticism (*what it is not*) (Stoltz & Ashby, 2007), its defining trait might have more to do with feeling satisfied and motivated while striving for high standards (*what it is*). Hence, the beneficial effects of self-compassion might be weaker in such a context, as suggested by a near-significant result (*p* = .07).

Self-compassion buffering the relationship between maladaptive but not adaptive perfectionism further supports the interpretation that self-acceptance might not be so integral to the trait of adaptive perfectionism. It also suggests that the nature of relationships between the two dimensions of perfectionism with self-esteem might be different. More precisely, the relationship between maladaptive perfectionism and lowered self-esteem might be stronger due to a lack of self-compassion (e.g., harsh self-criticism, feeling alone and inadequate, ruminating about one’s inadequacies). However, the relationship between adaptive perfectionism and increased self-esteem is not strengthened by being more self-compassionate. This could imply that the component within adaptive perfectionism that is linked with higher self-esteem might have less to do with being self-accepting or mindful during setbacks, since, if so, having more self-compassion should enhance its relationship with increased self-esteem. Perhaps what makes adaptive perfectionism linked with higher self-esteem is simply due to individuals’ satisfaction and good feelings about themselves upon reaching high standards (Missildine, 1963). The self-accepting nature during setbacks as suggested in theories has less to do with increasing these individuals’ self-esteem.

Nevertheless, this could also reflect the limitation of operationalizing adaptive perfectionism. Existing scales, including the APS-R only capture the defining feature of striving for high standards, yet fail to also incorporate its self-accepting and flexible traits. Structural equation analyses indicate that high standards alone, in fact, is a component of a latent construct interpreted as self-punitiveness (Hull et al.,1991). With such measurement limitations, the current study might have failed to show adaptive perfectionism’s interaction with self-compassion. This could also be the reason why adaptive perfectionism did not show any relationship with psychological distress. Similarly, the limitation of operationalization could explain why ‘adaptive’ perfectionism was found in some studies to be linked with low self-acceptance and a conditional view of the self-contingent on meeting high standards (Flett et al., 1994; Flett et al., 2003). Lastly, a closer look at the literature reflects that most theorists speak of ‘adaptive perfectionists’—a type of individual, rather than ‘adaptive perfectionism’—a personality trait. In these conceptualizations, adaptive perfectionists exhibit multiple traits (e.g., feeling satisfied and showing flexibility) which are believed to be, and clinically observed to give rise to, positive psychological outcomes (e.g., Hamachek, 1978). However, mainstream measurements fail to capture these traits fully (i.e., most scales only captured the trait of striving for high standards). This could potentially explain why there is a discrepancy between theory and empirical evidence with regards to adaptive perfectionism.

#### Self-Compassion as a Moderator Between Adaptive Perfectionism and Psychological Distress

Self-compassion did not moderate the relationship between adaptive perfectionism and psychological distress. Since self-compassion also did not moderate the maladaptive dimension– psychological distress link, the same potential reason regarding the use of a composite scale of psychological distress applies. Still, it is also possible that self-compassion, again, is less related to the construct of adaptive perfectionism, or that the self-accepting nature of adaptive perfectionism had not been captured due to the limitation with measurement. Moreover, self-compassion might also only enhance adaptive perfectionism’s relationship with *positive* well-being, rather than psychological distress (Brenner et al., 2018).

### Bidimensional Perfectionism

In this study, maladaptive perfectionism was found to be linked with higher psychological distress, whilst adaptive perfectionism did not show any relationship with psychological distress. This is in keeping with the bidimensional view of perfectionism (Gäde et al., 2017). In particular, although adaptive perfectionism did not predict lower psychological distress, its ‘adaptiveness’ might lie in its ability to give rise to positive psychological outcomes (Stoeber & Otto, 2006), for instance, increased self-esteem in this study.

Mediation results also provided evidence for the integral role of self-esteem in both dimensions of perfectionism (Preusser et al., 1994). A lowered self-esteem could explain why maladaptive perfectionism is linked with higher psychological distress, whereby an increase in self-esteem is one of the pathways through which adaptive perfectionism is linked with lower psychological distress. Whilst maladaptive perfectionism showed a clearer relationship with psychological distress via self-esteem, results on adaptive perfectionism suggest that it is a much more complicated construct. Though linked with higher self-esteem and in turn lowered psychological distress, adaptive perfectionism could give rise to other negative outcomes, as reflected by an insignificant total effect. This has a few implications. First, adaptive perfectionism might not be so adaptive in some aspects. For instance, striving for high standards might just be a manifestation of conditional self-acceptance, with this argument already existing in the current literature (Flett & Hewitt, 2002; Greenspan, 2000). However, this could also reflect the limitation of operationalizing adaptive perfectionism. Existing scales, including the APS-R only capture the defining feature of striving for high standards, yet fail to also incorporate its self-accepting and flexible traits. Adaptive perfectionism might not have been properly operationalized all along in our literature, leading to discrepancies between theory and empirical evidence. Further exploration with a more accurate operationalization of adaptive perfectionism is needed to understand whether adaptive perfectionism is indeed adaptive, or whether it might just be merely a reflection of conscientiousness or achievement striving, as argued by some scholars (Flett & Hewitt, 2006; Greenspan, 2000).

Self-compassion was found to buffer the positive link between maladaptive perfectionism and self-esteem. This suggests that showing self-kindness, being mindful, and recognizing common humanity during setbacks attenuates maladaptive perfectionism threat to lowered self-esteem. However, self-compassion did not enhance adaptive perfectionism’s effect on increased self-esteem. This further suggests that *maladaptive and adaptive perfectionism might not be flip sides of the same coin*—the mechanisms through which maladaptive and adaptive perfectionism leads to lowered/increased self-esteem might be different. The critical and ruminative ways of relating to oneself might be what contributes to lowered self-esteem associated with maladaptive perfectionism; whilst it is a different nature of adaptive perfectionism (not necessarily the lack of self-criticism) that contributes to its links with increased self-esteem.

### Strengths, Limitations, and Future Research

Although perfectionism is a well-researched topic, results regarding its two dimension’s relation with psychological distress remain mixed. By adopting multiple modes of statistical analysis—correlational, mediational, and moderation, whilst studying the two dimensions simultaneously, this study provides greater insights into the intricate mechanisms of bidimensional perfectionism, enabling us to understand it from a vantage point. Another major strength of this study lies in its addressing of the two major limitations of previous studies—the use of impure measurements and not partialling the two dimensions’ effects on one another. As reflected in this study, addressing these limitations impacts the observed relationships.

Nevertheless, this study still comes with some limitations. First, despite being a purer scale, the APS-R, like other major perfectionism scales, does not fully operationalize the self-accepting and flexible nature of adaptive perfectionism. Scholars should strive to develop a measurement of perfectionism that operationalize adaptive perfectionism more fully. Second, although partialling allows us to ‘understand the shared, unique, combined, and interactive relations of the [two] dimensions of perfectionism’ (Stoeber & Gaudreau, 2017, p.1), individuals are likely to exhibit both dimensions simultaneously, as reflected by their significant correlation.

This may cause the dimensions to manifest differently amongst different individuals in real life, reducing the generalizability of this study’s results. Future studies could replicate and extend this study by adopting cluster analysis to identify types of perfectionists. Thirdly, the use of a composite measure of psychological distress has limited the exploration of bidimensional perfectionism’s relationship with depressive and anxiety symptoms. Depression and anxiety are distinct phenomena (Clark & de Silva, 1985). As explored, the inability to replicate previously found interaction effect between maladaptive perfectionism and self-compassion on psychological distress could also be attributed to this reason. Future studies on perfectionism should measure anxiety and depressive symptoms separately as distinct psychological outcomes, discovering their unique relationships with self-esteem and self-compassion.

Other general limitations also apply. First, convenient sampling limits the generalizability of results. Random sampling should be conducted in the future so that the population is more accurately represented. Second, a cross-sectional design means no causality and directionality could be drawn. Although the current study suggests that perfectionism is linked to psychological distress via self-esteem, theorists like Hollender (1965) suggest that lowered self-esteem is not the result, but the cause of perfectionism. To better understand the relationships amongst the constructs in question, longitudinal and experimental designs should be adopted in the future. Lastly, lengthy online surveys might have caused fatigue and response bias amongst participants. This is especially true for the SCS which consists of 26 similar-worded questions.

Participants might not have paid close attention and responded carelessly. The validated SCS-short form (Raes et al., 2011) could be used in the future, although one should be aware that this form is less reliable if the subscale scores are of study interest (Raes et al., 2011).

### Practical Implications

Since only maladaptive perfectionism was linked to psychological distress, more emphasis should be put on this dimension in interventions. In particular, given the mediating role of self-esteem, clinicians should be aware of how self-esteem underpins the psychological and behavioral processes of perfectionistic individuals.

Other than working on maladaptive perfectionistic tendencies that may contribute to lowered self-esteem, self-esteem enhancing interventions could be done in tandem. Self-esteem interventions that are currently found effective include CBT, reminiscence-based interventions, and evaluative conditioning (Niveau et al., 2021). Still, their effectiveness in the context of perfectionism has not been studied, and the current research sets the stage for these research in the future.

Moreover, since self-compassion was found to buffer maladaptive perfectionism’s threat on lowered self-esteem, self-compassion interventions may also benefit perfectionist individuals. Self-compassion interventions were found to be effective for various psychological outcomes (Ferrari et al., 2019), and its benefits on self-esteem and perfectionism warrants further research. Nevertheless, it is important to note that the moderating effect of self-compassion remains small in this study, which might limit the cost-effectiveness of real-life interventions. This could be attributed to trait, rather than state self-compassion being measured (Faustino, 2022). State self-compassion might more effectively buffer the moment-to-moment threats of maladaptive perfectionism on lowered self-esteem, and such distinction should also be tackled in future interventional research.

## Conclusion

Through adopting a comprehensive study design: the use of correlational, mediational, and moderation analysis whilst studying both dimensions of perfectionism simultaneously, the current study shed light on the mixed literature regarding bidimensional perfectionism and psychological distress. The current study is in keeping with the bidimensional view of perfectionism, showing that only the maladaptive but not the adaptive dimension of perfectionism is related to psychological distress. Self-esteem was also found to mediate both dimensions’ relationship with psychological distress, suggesting that the idea of self-worth is integral to both dimensions of perfectionism. However, the current study reflects that maladaptive and adaptive perfectionism are not flip sides of the same coin. Whilst maladaptive perfectionism showed a more direct relationship with psychological distress, adaptive perfectionism may positively relate to psychological distress via some other pathways (e.g., conditional self-acceptance), despite showing a negative relationship with psychological distress via increased self-esteem. This could be due to adaptive perfectionism being more innately related to striving for achievements, rather than showing self-acceptance during setbacks, as reflected by a lack of interaction with self-compassion. It could also be due to the limitation in its measurement that fails to capture its self-accepting nature. Although APS-R was suggested to be a purer scale, future research should aim at developing a more accurate operationalization of adaptive perfectionism as suggested in theories. Moreover, potential negative mediators (e.g., conditional self-acceptance, social comparison) could be explored together with self-esteem, so as to better understand the ostensibly lack of relationship between adaptive perfectionism and psychological distress.

## Data Availability

All data produced in the present study are available upon reasonable request to the authors.

## Appendices

## Appendix A Informed Consent Forms

### Informed Consent Form

You are invited to participate in a research study conducted by Ms. Peony Chung under the supervision of Dr. Antoinette Lee from the Department of Psychology at the University of Hong Kong.

#### PURPOSE OF THE STUDY

The purpose of this study is to investigate the relationships among perfectionism, self-compassion, self-esteem, and psychological distress.

#### PROCEDURES

You will be invited to fill out an anonymized survey. It should take around 5-8 minutes.

#### POTENTIAL RISKS / DISCOMFORTS AND THEIR MINIMIZATION

This procedure has no known risks, nor should it pose any discomfort greater than what we experience in daily life. However, if you experience any discomfort at any point in this study, you are encouraged to take a break or withdraw if needed.

#### COMPENSATION FOR PARTICIPATION

Participations will be voluntary and no compensation will be provided.

#### POTENTIAL BENEFITS

There will be no direct benefit to you from participating in this study. However, your participation can help shed light on the topics of perfectionism, self-compassion, and self-esteem.

#### CONFIDENTIALITY AND DATA STORAGE

Your identity will be kept anonymous. Data collected will be kept strictly confidential and will be used for research purposes only. Only the primary investigator and supervisor will have access to these data, with these data being stored in a secure location only known to the primary investigator.

#### PARTICIPATION AND WITHDRAWAL

Your participation is voluntary. This means that you can choose to stop at any time without negative consequences.

#### QUESTIONS AND CONCERNS

If you have any questions about the research, please feel free to contact Ms. Peony Chung or Dr. Antoinette Lee. If you have questions about your rights as a research participant, contact the Departmental Research Ethics Committee, Department of Psychology, HKU (3917-5867).

**I understand the procedures described above and agree to participate in this study.**

## Appendix B Demographic Questions

1. Please indicate your age:
2. Gender pronouns:

- She/Her
- He/Him
- They/them
- Prefer not to say
- Other (Please specify)
3. Are you currently a student?

- Yes
- No
4. Please indicate your current level of education:

- Primary or below
- Secondary
- Higher/ Professional Diploma
- Bachelor’s degree
- Master’s degree
- Doctorate degree
- Other (Please specify)
5. Please indicate your family monthly income:

- Less than 5,000 HKD
- 5,000 - 10,000 HKD
- 10,001 - 20,000 HKD
- 20,001 - 40,000 HKD
- 40,001 - 80,000 HKD
- 80,001 - 150,000 HKD
- More than 150,000 HKD
6. Please indicate your ethnicity:

- Asian
- White
- American Indian
- Black or African American
- Hispanic Latino or Spanish origin
- Middle Eastern or North African
- Others (Please specify:)

## Appendix C Almost Perfect Scaled-Revised (APS-R) (‘Discrepancy’ and ‘High Standards’ Subscales) (Slaney et al., 2001)

***Note.*** Questions were presented in a randomized order.

Please indicate your degree of agreement with each of the following item. There are no right or wrong answers. Please use your first impression and do not spend too much time on individual items. (1 = Strongly Disagree, 2 = Disagree, 3 = Slightly Disagree, 4 = Neutral, 5 = Slightly Agree, 6 = Agree, 7 = Strongly Agree)

### Discrepancy

1. I often feel frustrated because I can’t meet my goals.
2. My best just never seems to be good enough for me.
3. I rarely live up to my high standards.
4. Doing my best never seems to be enough.
5. I am never satisfied with my accomplishments.
6. I often worry about not measuring up to my own expectations.
7. My performance rarely measures up to my standards.
8. I am not satisfied even when I know I have done my best.
9. I am seldom able to meet my own high standards of performance.
10. I am hardly ever satisfied with my performance.
11. I hardly ever feel that what I’ve done is good enough.
12. I often feel disappointment after completing a task because I know I could have done better.

### High Standards

1. I have high standards for my performance at work or at school.
2. If you don’t expect much out of yourself, you will never succeed.
3. I have high expectations for myself.
4. I set very high standards for myself.
5. I expect the best from myself.
6. I try to do my best at everything I do.
7. I have a strong need to strive for excellence.

## Appendix D Rosenberg Self-Esteem Scale (RSES) (Rosenberg, 1965)

Please indicate your degree of agreement with each of the following item. (1 = Strongly agree, 2 = Agree, 3 = Disagree, 4 = Strongly disagree)

1. On the whole, I am satisfied with myself.
2. At times I think I am no good at all.
3. I feel that I have a number of good qualities.
4. I am able to do things as well as most other people.
5. I feel I do not have much to be proud of.
6. I certainly feel useless at times.
7. I feel that I’m a person of worth, at least on an equal plane with others.
8. I wish I could have more respect for myself.
9. All in all, I am inclined to think that I am a failure.
10. I take a positive attitude toward myself.

## Appendix E Self-Compassion Scale (SCS) (Neff, 2003)

Please indicate how often you behave in the stated manner, using the following 1-5 scale. (1= Almost Never, 5 = Always)

1. I’m disapproving and judgmental about my own flaws and inadequacies.
2. When I’m feeling down I tend to obsess and fixate on everything that’s wrong.
3. When things are going badly for me, I see the difficulties as part of life that everyone goes through.
4. When I think about my inadequacies, it tends to make me feel more separate and cut off from the rest of the world.
5. I try to be loving towards myself when I’m feeling emotional pain.
6. When I fail at something important to me I become consumed by feelings of inadequacy.
7. When I’m down, I remind myself that there are lots of other people in the world feeling like I am.
8. When times are really difficult, I tend to be tough on myself.
9. When something upsets me I try to keep my emotions in balance.
10. When I feel inadequate in some way, I try to remind myself that feelings of inadequacy are shared by most people.
11. I’m intolerant and impatient towards those aspects of my personality I don’t like.
12. When I’m going through a very hard time, I give myself the caring and tenderness I need.
13. When I’m feeling down, I tend to feel like most other people are probably happier than I am.
14. When something painful happens I try to take a balanced view of the situation.
15. I try to see my failings as part of the human condition.
16. When I see aspects of myself that I don’t like, I get down on myself.
17. When I fail at something important to me I try to keep things in perspective.
18. When I’m really struggling, I tend to feel like other people must be having an easier time of it.
19. I’m kind to myself when I’m experiencing suffering.
20. When something upsets me I get carried away with my feelings
21. I can be a bit cold-hearted towards myself when I’m experiencing suffering
22. When I’m feeling down I try to approach my feelings with curiosity and openness
23. I’m tolerant of my own flaws and inadequacies
24. When something painful happens I tend to blow the incident out of proportion
25. When I fail at something that’s important to me, I tend to feel alone in my failure
26. I try to be understanding and patient towards those aspects of my personality I don’t like

## Appendix F Kessler Psychological Distress Scale (K10) (Kessler, 1996)

### In the past four weeks, about how often did you feel

1. tired out for no good reason?
2. nervous?
3. so nervous that nothing could calm you down?
4. hopeless?
5. restless or fidgety?
6. so restless you could not sit still?
7. depressed?
8. that everything is an effort?
9. so sad that nothing could cheer you up?
10. worthless?

(Options: ‘None of the time’, ‘A little of the time’, ‘Some of the time’, ‘Most of the time’, ‘All of the time’)

## Appendix D Debriefing Notes

### Debriefing Notes

Perfectionism as a personality trait can be seen as having both adaptive and maladaptive dimensions: Adaptive perfectionism is linked with the ability to derive satisfaction from striving for high standards whilst demonstrating flexibility and self-acceptance in face of mistakes.

Meanwhile, maladaptive perfectionism is linked with tendencies of harsh self-criticism and dissatisfaction when striving for unrealistically high standards (Stoeber, 2017).

Nevertheless, their relationships with psychological distress remains unclear in the current literature, warranting further research (Hewitt & Flett, 1991). In particular, self-esteem, the general attitude and evaluation of oneself (Rosenberg, 1965), and self-compassion, which entails having an open, kind, and nonjudgmental attitude towards one’s sufferings and shortcomings (Neff, 2003), are two traits that are believed to underlie the relationships between perfectionism and its psychological outcomes. This will be the first study to elucidate these relationships, and it is hoped that it could contribute to our current understanding of perfectionism. It is hypothesized that higher self-compassion could enhance the effects of adaptive perfectionism on increased self-esteem and reduced psychological distress; whereby it could buffer the effects of maladaptive perfectionism on reduced self-esteem and enhanced psychological distress.

Thank you for participating in this research. If you have any questions about the research, please feel free to contact Ms. Peony Chung or Dr. Antoinette Lee. This study has been reviewed by, and received human research ethics approval from the Departmental Research Ethics Committee, Department of Psychology, The University of Hong Kong. If you have questions about the research ethics approval of this project, please contact the General Office of the Department of Psychology, HKU (3917-5867).

## Notes

### Competing Interest Statement

The authors have declared no competing interest.

### Funding Statement

This study did not receive any funding

### Author Declarations

This study has been reviewed by, and received human research ethics approval from the Departmental Research Ethics Committee, Department of Psychology, The University of Hong Kong.

